# Determining latent features and forecasting of COVID-19 hospitalisations in Malaysia using a national patient assessment data platform: a study of machine learning modelling against expert system

**DOI:** 10.1101/2023.01.17.22281858

**Authors:** Hui-Jia Yee, Ivan Boo, Ian K.T. Tan, Jih Soong Tan, Helmi Zakariah

## Abstract

COVID-19 had a severe impact on Malaysia, as cases increased dramatically as the pandemic spread. In order to combat the pandemic, the Ministry of Health has established a number of standard operating procedures (SOP) and started operating COVID-19 Assessment Centers (CAC). This study compares the expert system created using the current patient evaluation standards to the capabilities of machine learning approaches in capturing the potential of being admitted directly or during home quarantine, based on the different clinical symptoms and age group. Boruta is a feature selection method that is employed to rank and extract significant characteristics.

Treatment for imbalance has been carried out by under-sampling with K-Means and over-sampling with SMOTE. It appeared that the machine learning method using Random Forest would perform better than the expert systems. There are five performance metrics used in this study, i.e. accuracy, precision, recall, F1-score, and specificity. This study focused to maximize the true positive rate while minimize the false negative rates, it is to make sure that the patient who really need to be hospitalized will not be missed out. Therefore, recall becomes the main evaluation metrics when comparing the machine learning model and the expert system. The results shown that the recall score for machine learning approach is vastly higher then of expert systems. For age group 18-59, machine learning has 32.75% recall more than the expert system to predict if a patient requires direct admission, while for age group more than 60, the recall of machine learning is 18.11% more than expert system. In addition, to predict if a patient require admission during their home quarantine due to their health deterioration, machine learning recorded 76.72% recall more than the expert system for patient aged 18 to 59, and 70.59% difference for patient more than 60 years old. This supports the potential application of machine learning for clinical decision making for COVID-19 patients.

## Introduction

Coronavirus disease 2019 (COVID-19) pandemic is the greatest communicable disease outbreak to have hit Malaysia [1]. In Malaysia, the number of deaths began to rise on February 7, 2022, reaching a peak of 89 on March 14, 2022, while Malaysia reached 32800 cases on March 10, 2022 [2]. COVID-19 is caused by severe acute respiratory syndrome coronavirus 2 (SARS-CoV-2) and is transmitted by respiratory droplets and contacts [3]. The main manifestations of COVID-19 include fever and dry cough and most COVID-19 cases are non-severe with favourable outcomes [4, 5].

To protect the health of Malaysian citizens, several actions were taken by the Malaysian government. These include the enforcement of health screening at all entry points, increasing the number of hospitals that could treat COVID-19 cases, and restrictions on people mobility through the Movement Control Order (MCO) which was implemented on March 18, 2020 [6]. In addition, as part of Ministry of Health (MOH)’s effort to counter COVID-19 spread, they utilised convention centres, public halls, and indoor stadiums as quarantine and treatment centres for low-risk patients [6].

As the number of patients outgrew the capacity of these treatment centres, and with better understanding of the spread severity of the disease, home quarantine was allowed for patients who are asymptomatic or have very mild symptoms. With this, Malaysia’s MOH set up 213 COVID-19 Assessment Centers (CAC) nationwide. The main function of CAC is to conduct assessments of COVID-19 patients undergoing treatment at home [7]. CAC help the medical front-liners to monitor patients’ condition and determine if they either need to be home quarantined or hospitalised. In Malaysia, the doctors at the CACs follow a series of prerequisite guidelines that would determine whether the patient is allowed to be home quarantined or requires direct admission into hospitals [8].

The CACs had assessed hundreds of thousands of patients and the data collected enabled our study on the effectiveness of using machine learning (ML) vis-a-vis assessment guidelines for determining whether a patient needs to be admitted and also whether a home quarantined patient’s health will deteriorate to a level that requires hospital admission. This is an important study as with the pandemic moving towards an endemic phase, Malaysia has shut down the physical CACs but will continue with virtual CACs that conducts assessment online. Therefore, the objectives of this study are:

1. To identify the predictive factors in order to determine whether a patient requires direct admission or the patient can be monitored under home quarantine.
2. To predict if a home quarantine patient requires admission during their home quarantine, due to the deterioration of their health status.

These are evaluated by comparing the machine learning method with an expert system that is built based on the existing patient assessment guidelines. A random guessing method is also included in the evaluation as an indicative measure of the effectiveness of the expert system.

### Machine Learning Challenges in Medical Diagnosis

Based on latent patterns throughout massive datasets, machine learning can be used to comprehend, predict, and steer future behaviour. This allows for better decisions to be made based on different context with the data mining process [9].

### Machine Learning Algorithm for COVID-19

Random Forest (RF), and ensemble-based ML algorithms, has been applied to predict the severity and outcome of COVID-19 patients [10–15]. Using a collection of decision trees, RF, a type of machine learning, can assess intricate relationships between clinical traits and offer highly accurate classifications [16]. To further help hospitals and medical facilities decide on prioritorising patients for hospitalisation, RF have been implemented [17] with features selected using the filter and wrapper method. The feature selection method is used to reduce the dimensionality and complexity by filtering the most informative feature and eliminating redundant data. RF was also applied to a similar problem setting in predicting the ICU admission rate of COVID-19 patients [18]. It exhibits the best performance among other well-known tree-type and ensemble algorithms. These studies provided convincing grounds for applying ML, specifically RF, in order to achieve our objectives.

### Data Quality and Features of Medical Records

Dirty data plays a significant challenge when applying ML. Bellman’s Curse of Dimensionality stated that datasets with higher dimensionality succumb to the presence of dirty data [19], hence feature selection (FS) methods became an important practical area for researchers and practitioners alike. Conducting feature selection using the Boruta method yields an ordered list (ranking) of features based on their importance [20, 21]. This method would eventually provide great insights to achieve the objective in determining the predictive factors for hospitalization. On the other hand, Boruta is available as an efficient implementation, making analysis of high-dimensional data sets feasible [20]. Nevertheless, a problem arises when given data sets are imbalanced in nature and the common technique to oversample a data set used in a typical classification problem is Synthetic Minority Over-sampling Technique (SMOTE) [21]. As observed from [16] where RF is used, together with the application of SMOTE and RBF as the feature selection method, allows the researcher to produce a high-performing model that could predict the clinical prognosis and outcome of hospitalized COVID-19 positive patients with the AUC Score of *≈* 99%.

### Imbalance Medical Records

To elaborate on the imbalance dataset, the scenario occurs when one or more classes (i.e., minority classes) have very few cases while other classes (i.e., majority classes) have large numbers of cases which causes the class distribution to be highly skewed in real-world applications [22]. For cases such as medical diagnoses the minority classes deserve special attention because it represents the phenomenon where the predicted outcome is among a plethora of majority class patterns denoting normal operations [23]. Brandt and Lanzén [24] shows that using SMOTE improves the relative performance of RF using imbalanced data sets. SMOTE uses the oversampling approach in which the key idea of SMOTE is to create new data based on the interpolation between several minority classes within a defined neighbourhood to re-balance the data set [25].

Additionally, SMOTE performed statistically significantly better on mammography data and several others, laying the groundwork for learning from imbalanced data sets [25].

The next section will elaborate on the source of data for our work, details on the pre-processing of the data for feature selection, our application of SMOTE on the imbalanced dataset, and the methods used to to the classifications, namely the expert system and the machine learning process.

## Methods

### Study Design and the Source of Data

SELangkah is a COVID-19 mobile app launched by the Selangor state government in May 2020 [26]. The features in the app covered the FFTISV (Find, Test, Trace, Isolate, Support and Vaccinate) strategy, which it was not only used for contact tracing, but also included the program for the people to register and go through community screening and vaccination. Besides, it implemented a home assessment tool for the medical officer to monitor patients under home quarantine. In collaboration with SELangkah, COVID-19 Assessment Centres (CAC) [27] offers datasets containing comprehensive patient data. The datasets offered comprise a wide range of valuable features that pique the attention of this study, including biological features, underlying comorbidities, symptoms, and triage categorization. The study is driven in the direction of the objectives by the availability of the clinical decision results (whether to be admitted directly or to be placed into home quarantine) as well as the outcome of home quarantine.

### Data Preparation

The data used in this study is between February 22, 2021 and April 13, 2022. In this empirical study, several sets of data cleaning procedures have been done, mainly from different approaches. These approaches are categorised and summarised into two approaches based on the main differences as in Table 1.

**Table 1.**
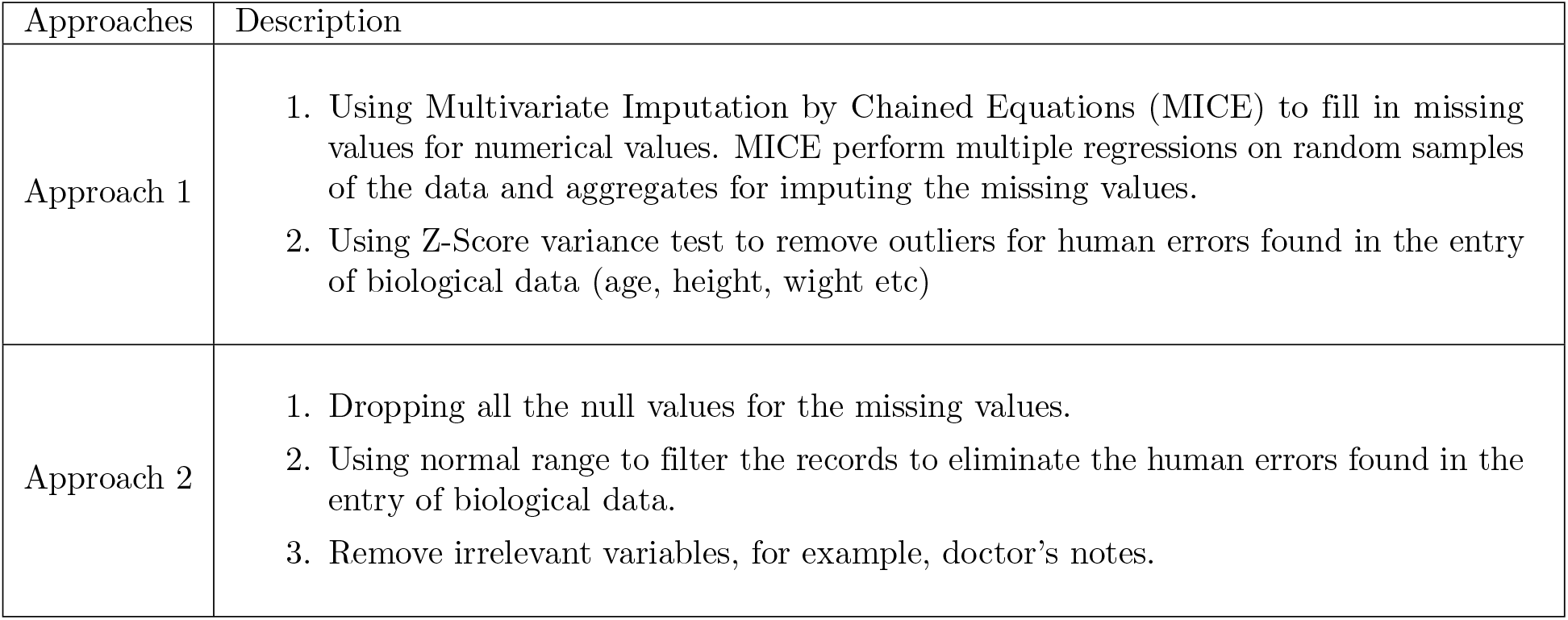
Data Cleaning Approaches.

In the initial phase, Approach 1 has been used to wrangle the analytical dataset for two reasons:

1. To prevent the loss of excessive records from the analytical datasets.
2. To remove records based on the variances as a proof for determining the correct range of which numerical range made sense.

However, with further consideration, imputation presents probable chances of introducing bias as well as artificial data that are deemed as noise to the dataset as most of the numerical variables that are having missing values constituted to ≈ 27% of the entire dataset. In addition, by using a repetitive Z-score variance to remove the outliers for human errors further remove additional records of the dataset, which it contradicts to the objective of reducing records due to missing values. Besides, the processes are difficult to be generalised in the future, as well as to make it more robust. Therefore, approach 2 has been proposed and implemented.

The main reason of applying approach 2 is to produce a “clean and pure” dataset that is free from noise and missing values. Even though the downside of this particular approach removes a huge chunk of records from the dataset, the properties of having a clean dataset is an advantage in the subsequent processes. After a clean dataset is produced, label encoding is performed for the categorical variables, which each label is assigned a unique integer.

Table 2 shows the number of rows and columns before and after performing data cleaning.

**Table 2.**
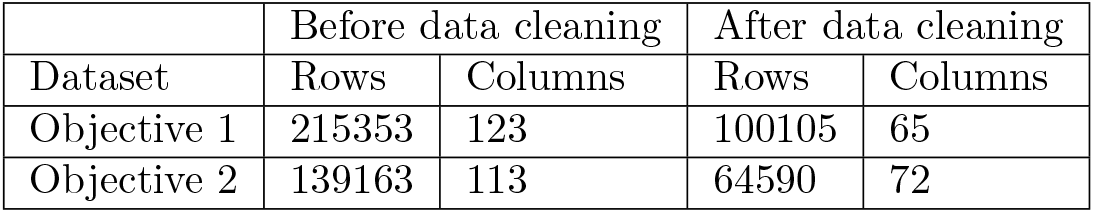
Comparison of Data Dimension Before and After Data Cleaning.

|  | Before data cleaning |  | After data cleaning |  |
| --- | --- | --- | --- | --- |
| Dataset | Rows | Columns | Rows | Columns |
| Objective 1 | 215353 | 123 | 100105 | 65 |
| Objective 2 | 139163 | 113 | 64590 | 72 |

### Feature Engineering

#### Data Splitting

Based on subject matter expert’s opinion, different age group of people has different impact from COVID-19 and a study by Zhang et. al [28] also shown that age is a risk factor for COVID-19 patients. Therefore, in this study, the dataset is split into 4 age groups,

1. ages 0-5,
2. ages 6-17,
3. ages 18-59, and
4. ages 60+.

After splitting the dataset, the total size of each of the age group dataset is analyzed. and (2) contains limited records, hence we are are unable to proceed. The remaining datasets, (3) and (4), are then split for training and testing with the ratio of 80:20; 80% of the data is used as the training data for feature selection and building models while the rest 20% is used as the test set for model validation purposes.

#### Feature Selection

After having the dataset split to two sets of data ((3) ages 18-59, (4) ages 60+), Boruta algorithm was used to perform feature selection on both the datasets.

Boruta is an all-relevant feature selection wrapper algorithm, it can be used with any classification methods to produce the variable importance measure (VIM) [29]. In this study, the classification method, Random Forest is used for Boruta. Boruta iteratively compares the original features’ importance with the importance of permuted features’ (shadow attributes) to remove the irrelevant features, which are the features having significantly less importance as compared to the permuted features. On the other hand, the features that are having significantly more importance than the shadow attributes will be confirmed as relevant features. In this study, 100 iterations were used to run the Boruta process. The feature selection algorithm will select a set of features that are of the most importance. The features selected for each objective are shown in Table 4 and Table 5.

#### Handling of Imbalanced Data

As expected, medical diagnosis datasets are imbalance in that there will be an overwhelming case of negative labels as oppose to those that are positive. Machine learning algorithms work best when the dataset is balanced in order not to have the predicted outcome to simple be part of the majority class which will then denote a high accuracy due to the normalcy of the classification. One of the ways to overcome this issue is by using the appropriate sampling methods. There are under-sampling methods and oversampling methods. In this study, SMOTE (Synthetic Minority Oversampling Technique) is used to perform oversampling to increase the samples of minority class while clustering approach is used to perform under-sampling to decrease the samples of majority class.

Clustering algorithm is implemented as an alternative way of treating the imbalance issue by under-sampling. The purpose of implementing this approach is to achieve a reduced sample size of majority class that contains the best general representation across the majority class.

Firstly, the dataset is separated into minority and majority class. In this case, most of the datasets are having ‘Negative’ class label as the majority class and ‘Positive’ as the minority class. Further on, a targeted size of majority class will be computed based on the number of minority class. A balancing ratio of 2:1 in terms of majority to minority class is applied in this exercise to allow oversampling to be performed later on. As an example, if there are 1000 records available for minority class, the targeted records for majority class will be 2000. This method is only applicable if the proportion of the majority class is two or more times larger than the minority class.

K-Means algorithm is first applied to the data of majority class, where the best number of clusters are determined by elbow method based on Distortion and Inertia. The proportion of the clusters is then calculated by computing the size of the clusters with the total records of majority class. As an example after the clustering process, 42% of the records categorized into Cluster 0, 40% are categorized into Cluster 1 and 18% categorized into of Cluster 2.

From each of the clusters, a sample of records will be extracted based on the proportion of computed target of records for majority class. If the targeted number of majority class is 2000, then the total records that needs to be subsampled from:

- Cluster 0 is 42% x 2000= 840
- Cluster 1 is 40% x 2000= 800
- Cluster 2 is 18% x 2000= 360

The total number of subsample should be almost the same as the targeted number of majority class. The extracted subsample of majority class data rows are then merged with the extracted minority class data rows to produce a dataset with the ratio of 2:1 by class. Subsequently, oversampling is performed to get a 1:1 balanced dataset and then the following process is continued as usual.

#### Model Building

In order to compare the performances of a medical professional using a set of guidelines with a machine learning model, an expert system representing the guidelines is also built alongside with the predictive (ML) model. Predictive (ML) models are constructed to forecast an output by leveraging the features that are selected by the feature selection algorithm, whereas expert systems replicate human judgement and follow specific rules provided in their knowledge base.

#### Expert System

The expert system (ES) is built with the purpose of simulating the diagnosis of doctors based on the guidelines set by Malaysia’s Ministry of Health [8]. Physicians are needed to adhere to the guidelines stated in Table 3 to determine if an adult COVID-19 patient is required to be admitted.

**Table 3.**
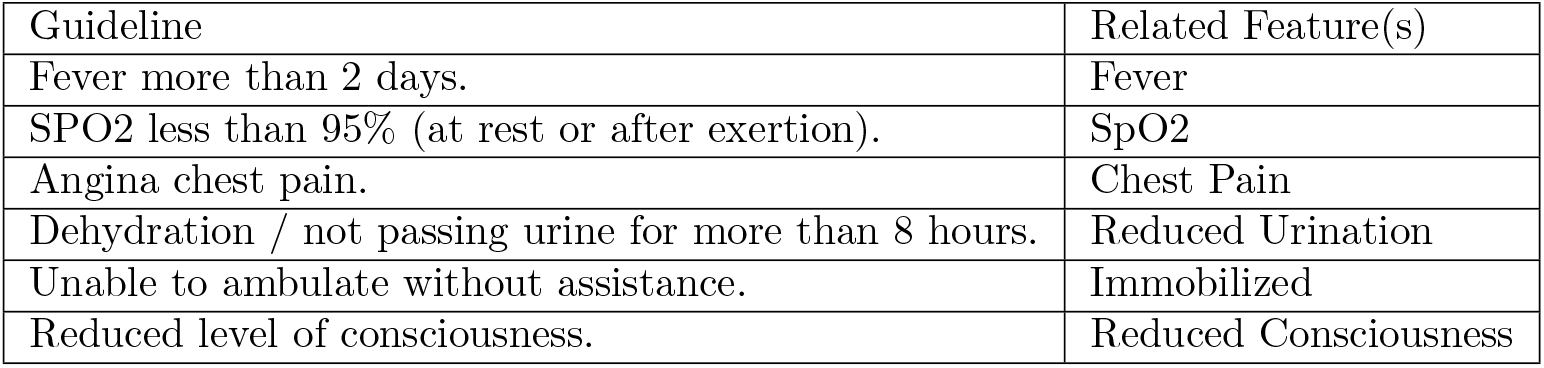
Guidelines for Admission for COVID-19 Patients.

| Guideline | Related Feature(s) |
| --- | --- |
| Fever more than 2 days. | Fever |
| SPO2 less than 95% (at rest or after exertion). | SpO2 |
| Angina chest pain. | Chest Pain |
| Dehydration / not passing urine for more than 8 hours. | Reduced Urination |
| Unable to ambulate without assistance. | Immobilized |
| Reduced level of consciousness. | Reduced Consciousness |

The suggested guidelines list the red flags that call for an admission of a COVID-19 patient. To build a deterministic output based on the rules generated from the recommendations in Table 3, a function has been developed. A patient would be considered for admission by the expert system if the patient meets any one of the guidelines’ cutoff points, mirroring the display of a doctor’s diagnosis for adult COVID-19 patients in Malaysia. Algorithm 1 describes the structure of the expert system.

##### Algorithm 1

Expert System Algorithm

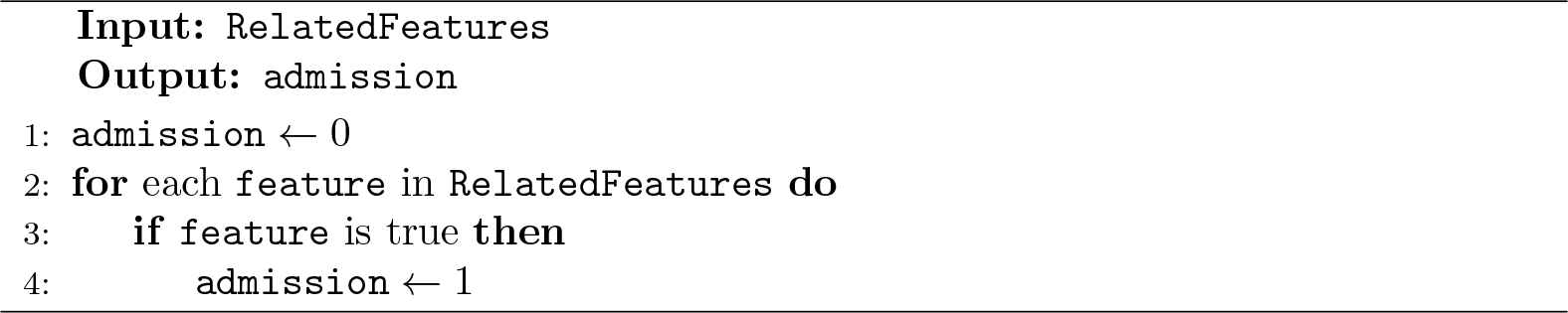

Algorithm 1 accepts a record with a list of related features that are fore-mentioned in Table 3 and transformed into a list `RelatedFeatures` as an input. The algorithm will set admission as a flag that indicates the need for admission to 0. The algorithm will further check if any of the features passed the threshold of the guidelines - where `admission` will be set to 1. The output of the algorithm returns the flag `admission` where:

- 0 - No admission required
- 1 - Admission required

#### Predictive Model

Random Forest (RF) is used as the classifier for model building. Random Forest is a supervised machine learning algorithm, it can be used for both classification and regression problems. RF introduces randomness into the model when it develops the decision tree, where the wide diversity of the decision trees created can construct a finer model. First, RF randomly select *n* features from a total *N* feature, it then split the node into two or more sub nodes using entropy or information gain calculation in order to identify the best split. The process is repeated until a maximum or user defined number of nodes is reached. These steps are used to construct a single tree. The forest (collection of trees) is built by repeating the steps of creation for single tree and then combine them. In other words, RF builds multiple decision trees and ensemble them to get a more accurate and stable prediction.

In addition, cross-validation (CV) and hyper-parameter tuning using Bayesian Optimisation [30] were performed. Hyperparameter-tuning is the process of searching the most accurate hyperparameters for a dataset with a ML algorithm. Bayesian Optimization runs models many times with different sets of hyperparameter values, but it evaluates the past model information to select hyperparameter values to build the newer model.

## Results

### Description

For objective 1, the final dataset included 58,327 COVID-19 patients aged 18 to 59 and 6617 patients more than 60 years old who visits CAC. On the other hand, for objective 2, the final dataset included 18,337 home quarantined patients aged 18 to 59 and 2670 patients aged 60 and above.

### Model predictors

The final features or predictors included in the models are shown in Table 4 for objective 1 and Table 5 for objective 2 with different age groups respectively.

**Table 4.** Feature Selected for Objective 1.

| Ages 18 -59 | Ages 60+ |
| --- | --- |
| Triage Category<sup>a</sup> | Triage Category<sup>a</sup> |
| Runny nose | Difficulty breathing |
| Cough | Fever |
| Difficulty breathing | Temperature |
| Fever | SpO<sub>2</sub><sup>b</sup> |
| Temperature | Lethargic |
| SpO<sub>2</sub><sup>b</sup> | Chest pain |
| Diarrhea | Excess lethargic |
| Vomit | Hypertension |
| Lethargic | Pulse rate |
| Myalgia | Respiratory rate |
| Chest pain | Weight |
| Loss appetite | Patient source<sup>c</sup> |
| Excess lethargic | Age |
| Worsening symptoms | Systolic blood pressure |
| Hypertension | Diastolic blood pressure |
| Diabetes Mellitus |  |
| Pregnancy |  |
| Pulse rate |  |
| Respiratory rate |  |
| Height |  |
| Weight |  |
| Patient source<sup>c</sup> |  |
| Gender |  |
| Race |  |
| Age |  |
| Systolic blood pressure |  |
| Diastolic blood pressure |  |

<sup>a</sup> Preliminary classification of patient based on symptom and their lung condition
<sup>b</sup> Oxygen saturation
<sup>c</sup> Origin of patient, e.g. hospital, district health office etc

**Table 5.** Feature Selected for Objective 2.

| Ages 18-59 | Ages 60+ |
| --- | --- |
| Triage Category<sup>a</sup> | Triage Category<sup>a</sup> |
| Runny nose | Runny nose |
| Cough | Cough |
| Temperature | Temperature |
| SpO<sub>2</sub><sup>b</sup> | SpO<sub>2</sub><sup>b</sup> |
| Loss of smell | Hypertension |
| Loss of taste | Pulse rate |
| Myalgia | Respiratory rate |
| Hypertension | Weight |
| Diabetes Mellitus | Patient source |
| Pregnancy | Race |
| Pulse rate | Age |
| Respiratory rate | Systolic blood pressure |
| Height | Diastolic blood pressure |
| Weight |  |
| Patient source<sup>c</sup> |  |
| Gender |  |
| Race |  |
| Age |  |
| Systolic blood pressure |  |
| Diastolic blood pressure |  |

<sup>a</sup> Preliminary classification of patient based on symptom and their lung condition
<sup>b</sup> Oxygen saturation
<sup>c</sup> Origin of patient, e.g. hospital, district health office etc

### Model evaluation

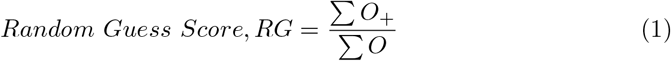

Let *O* represents the observations, or records in the dataset, *O*_+_ be the positive records. Equation 1 takes the quotient of total number of positives observations and the total number of all observation available. This equation is served as the benchmark for each model as it explains the general probability of a patient to be admitted based on the general probability of the dataset.

From the final datasets, 20% of the data is separated out to be used as the independent test set in order to evaluate the ML models. To preserve consistency, the expert system will be evaluated using the same test set. Table 6 (for Objective 1) and Table 7 (for Objective 2) compare the result of ML models and expert system. A comparison against the random guess (RG) score is also included in Table **??** and Table **??**.

**Table 6.** Objective 1 Result.

|  | Age 18-59 |  |  | Age 60 and more |  |  |
| --- | --- | --- | --- | --- | --- | --- |
|  | ML | ES | RG | ML | ES | RG |
| Accuracy | **83.11%** | 76.60% | 32.62%* | **76.28%** | 71.10% | 35.09%* |
| Precision | 78.15% | **82.58%** |  | **83.33%** | 78.99% |  |
| Recall | **68.57%** | 35.82% |  | **42.12%** | 24.01% |  |
| F1-Score | **73.05%** | 49.97% |  | **55.95%** | 36.82% |  |
| Specificity | 90.40% | **96.34%** |  | 95.31% | **96.55%** |  |

\* Random Guess Score

**Table 7.** Objective 2 Result.

|  | Age 18-59 |  |  | Age 60 and more |  |  |
| --- | --- | --- | --- | --- | --- | --- |
|  | ML | ES | RG | ML | ES | RG |
| Accuracy | **78.17%** | 66.93% | 31.90%* | **73.59%** | 66.75% | 32.08%* |
| Precision | **62.10%** | 35.09% |  | **56.98%** | 22.22% |  |
| Recall | **81.03%** | 4.31% |  | **72.06%** | 1.47% |  |
| F1-Score | **70.31%** | 7.68% |  | **63.64%** | 3.76% |  |
| Specificity | 76.83% | **96.26%** |  | 74.31% | **97.57%** |  |

\* Random Guess Score

## Discussion

Boruta is used as the feature selection algorithm to determine the predictive factors for hospital admission during visit CAC (Objective 1) or under home quarantine (Objective 2). The features selected or retained suggest that those features contribute most in predicting the hospitalization of COVID-19 patients.

As shown in Table 4 and Table 5, the features selected for age group more than 60 years old is a subset of the features selected for age group 18 to 59. Besides, the features *myalgia* (pain in a muscle or group of muscles) and *gender* are selected when predicting the hospitalization of patients aged 18 to 59 but not 60 or above. One of the reason inferred from this observation is, elderly people require less criteria to determine if they are to be admitted to hospital. There are 11 features that are always selected for each of the model in this study, namely, *triage category, temperature, SpO*_*2*_, *hypertension, pulse rate, respiratory rate, weight, patient source, age, systolic blood pressure* and *diastolic blood pressure*. Some the the variables can be easily accessible using a thermometer, pulse oximeter and sphygmomanometer, this study reveals the importance to monitor the vital signs of a patient to prevent further deterioration of health condition. As of 10 March 2022, the Malaysia government has distributed over 2 million COVID-19 care packages to the families in the bottom 40 per cent (B40) income category [31].

Among all the comorbidities *(hypertension, diabetes mellitus, illness related to nerve, liver, metabolic, blood, or immune system, lung disease and heart disease)* data collected, *hypertension* plays as one of the significant roles in determining whether a patient should be hospitalized, for all age groups. Yet, *diabetes* as a predictive factor only selected for age group 18 to 59 but not for the 60+ age group. Notably, COVID-19 as an infectious disease caused by the SARS-CoV-2 virus and will lead to mild to moderate respiratory illness, *lung disease* is not selected as a predictive factor in both age groups. By comparing the accuracy between ML, ES and Random Guessing based on Table Table 4 and Table 5, ML and ES is exhibiting a greater accuracy compared to the random guessing method. This scenario displayed that ML and ES is generally better in capturing the correct outcome of the patients compared to the dependence of the general probability of the dataset.

Based on Table 6 and Table 7, it clearly show that, the overall performance of ML models are better than the ES models. In this study, the objective is to predict whether a patient need to be hospitalized when they visit CAC or when they are under home quarantine with the eventuality of needing to be admitted, hence a high recall is crucial to make sure the patient who really need to be hospitalized will not be missed out. In other words, true positive rate needs to be increased whilst false negative rate needs to be decreased. On the account of that, this study focus on the recall to evaluate the different models and expert system. In term of recall, ML models outperformed significantly than the ES models for objective 1 and objective 2.

To predict the patients that needed to be hospitalized during their home quarantine (objective 2), the ES models for both age group recorded extremely low recall and f1-score as opposed to ML model as well as the random guessing score. In comparison of specificity and the sensitivity (recall) score, expert system is only able to capture the patients that will not be admitted, which is the true negative rate rather than the true positive rate. In other words, expert system is only performing well in classifying negative scenarios.

By comparing all the performance metrics (except specificity), ML models have higher accuracy, precision, recall and f1-score than the expert systems. Nonetheless, for objective 1 age group 18 to 59, the precision of expert system, which is 82.58% is higher than the ML model. It can be concluded that for expert system, there is one set of rule to follow in order to determine if patients need to be admitted to hospital when they visit a CAC, and hence it is able to ensure the consistency of the medical officer’s decision. On the contrary, the precision is low when the expert system is used to determine if a patient needs to be hospitalized under home quarantine. As patients are being remotely monitored during their home quarantine period, more variables or predictors need to be taken into consideration when examine their health conditions. The ES algorithm used in this study is the same for objective 1 and 2, this might lead to the deduction that the ES algorithm is not suitable to be used in determining a patient that needs to be hospitalized under home quarantine.

In a bigger picture, ES are predefined rules that are difficult to fixate to all scenarios across all population in Malaysia, as every set of population may contain unseen patterns and relationships in between the mined features that leads to a better analysis and prediction by applying machine learning models for its pattern mining properties.

## Data Availability

https://www.kaggle.com/datasets/yhuijia/covid19-hospitalization-data

## Acknowledgments

The authors would like to acknowledge the patients who registered in CAC, the doctors and nurses who cared for the patients, and the medical officers at the COVID-19 Assessment Centre.

## Notes

### Competing Interest Statement

The authors have declared no competing interest.

### Author Declarations

Professor Ruth Aylett of Mathematical and Computer Sciences Ethics Approval Team gave ethical approval for this work.

## References

1. Jayaraj VJ, Rampal S, Ng CW, Chong DWQ. The Epidemiology of COVID-19 in Malaysia. The Lancet Regional Health - Western Pacific. 2021;17:100295. doi:https://doi.org/10.1016/j.lanwpc.2021.100295.

2. COVIDNOW in Malaysia;. Available from: https://covidnow.moh.gov.my/.

3. Geng MJ, Wang LP, Ren X, Yu JX, Chang ZR, Zheng CJ, et al. Risk factors for developing severe COVID-19 in China: an analysis of disease surveillance data. Infectious Diseases of Poverty. 2021;10(1):48. doi:10.1186/s40249-021-00820-9.

4. Song J, Park DW, Cha Jh, Seok H, Kim JY, Park J, et al. Clinical course and risk factors of fatal adverse outcomes in COVID-19 patients in Korea: a nationwide retrospective cohort study. Scientific Reports. 2021;11(1):10066. doi:10.1038/s41598-021-89548-y.

5. Xiong Y, Ma Y, Ruan L, Li D, Lu C, Huang L, et al. Comparing different machine learning techniques for predicting COVID-19 severity. Infectious Diseases of Poverty. 2022;11(1):19. doi:10.1186/s40249-022-00946-4.

6. Shah AUM, Safri SNA, Thevadas R, Noordin NK, Rahman AA, Sekawi Z, et al. COVID-19 outbreak in Malaysia: Actions taken by the Malaysian government. International Journal of Infectious Diseases. 2020;97:108–116. doi:https://doi.org/10.1016/j.ijid.2020.05.093.

7. Ang J. Malaysia government sets up 213 COVID-19 assessment centers nationwide;. Available from: https://www.humanresourcesonline.net/malaysia-government-sets-up-213-covid-19-assessment-centers-nationwi

8. Guideline on home monitoring and management of confirmed COVID-19 case …;. Available from: https://covid-19.moh.gov.my/garis-panduan/garis-panduan-kkm/ANNEX-2m-Guideline-on-Home-Monitoring-n-Mgt-of-Confirmed-COVID-19-Capdf.

9. Jackins V, Vimal S, Kaliappan M, Lee MY. AI-based smart prediction of clinical disease using random forest classifier and naive bayes. The Journal of Supercomputing. 2020;77(5):5198–5219. doi:10.1007/s11227-020-03481-x.

10. Casiraghi E, Malchiodi D, Trucco G, Frasca M, Cappelletti L, Fontana T, et al. Explainable Machine Learning for Early Assessment of COVID-19 Risk Prediction in Emergency Departments. IEEE Access. 2020;8:196299–196325. doi:10.1109/ACCESS.2020.3034032.

11. Iwendi C, Bashir AK, Peshkar A, r S, Chatterjee JM, Pasupuleti S, et al. COVID-19 patient health prediction using boosted random forest algorithm. Frontiers in Public Health. 2020;8. doi:10.3389/fpubh.2020.00357.

12. afrash mr, Yaghoubi M, Rahimi F, Shanbehzadeh M, Bahadori M. An intelligent system for prediction of severity of SARS-COV-2 infection and progression to critical illness: Using machine learning models. 2021;doi:10.21203/rs.3.rs-1074101/v1.

13. Gök EC, Olgun MO. SMOTE-NC and gradient boosting imputation based random forest classifier for predicting severity level of covid-19 patients with blood samples. Neural Computing and Applications. 2021;33(22):15693–15707. doi:10.1007/s00521-021-06189-y.

14. Aljameel SS, Khan IU, Aslam N, Aljabri M, Alsulmi ES. Machine Learning-Based Model to Predict the Disease Severity and Outcome in COVID-19 Patients. Scientific Programming. 2021;2021:5587188. doi:10.1155/2021/5587188.

15. Laatifi M, Douzi S, Bouklouz A, Ezzine H, Jaafari J, Zaid Y, et al. Machine learning approaches in Covid-19 severity risk prediction in Morocco. Journal of Big Data. 2022;9(1):5. doi:10.1186/s40537-021-00557-0.

16. Wang J, Yu H, Hua Q, Jing S, Liu Z, Peng X, et al. A descriptive study of random forest algorithm for predicting COVID-19 patients outcome. PeerJ. 2020;8:e9945.

17. Pourhomayoun M, Shakibi M. Predicting mortality risk in patients with COVID-19 using machine learning to help medical decision-making. Smart Health. 2021;20:100178. doi:https://doi.org/10.1016/j.smhl.2020.100178.

18. Verma A, Patel AB, Hardin CC, Khandekar MJ, Lee H, Stylianopoulos T, et al. Comparing machine learning algorithms for predicting ICU admission and mortality in COVID-19. NPJ Digital Medicine. 2020;doi:10.1101/2020.11.20.20235598.

19. Venkat N. The curse of dimensionality: Inside out. Pilani (IN): Birla Institute of Technology and Science, Pilani, Department of Computer Science and Information Systems. 2018;.

20. Kumar SS, Shaikh T. Empirical Evaluation of the Performance of Feature Selection Approaches on Random Forest. In: 2017 International Conference on Computer and Applications (ICCA); 2017. p. 227–231.

21. Selvaraj C, Natarajan B, Kb S. Empirical study of feature selection methods over classification algorithms. International Journal of Intelligent Systems Technologies and Applications. 2018;17:98. doi:10.1504/IJISTA.2018.10012887.

22. A novel oversampling technique for class-imbalanced learning based on SMOTE and natural neighbors. Information Sciences. 2021;565:438–455. doi:https://doi.org/10.1016/j.ins.2021.03.041.

23. Elreedy D, Atiya AF. A Comprehensive Analysis of Synthetic Minority Oversampling Technique (SMOTE) for handling class imbalance. Information Sciences. 2019;505:32–64. doi:https://doi.org/10.1016/j.ins.2019.07.070.

24. Brandt J, Lanzén E. A comparative review of smote and ADASYN in Imbalanced Data Classification; 2021. Available from: http://www.diva-portal.org/smash/record.jsf?pid=diva2%3A1519153&dswid=-3233#:~:text=The%20results%20show%20that%20both,degree%20of%20class%20imbalance%20increases.

25. Fernandez A, Garcia S, Herrera F, Chawla NV. Smote for learning from imbalanced data: Progress and challenges, marking the 15-year anniversary. Journal of Artificial Intelligence Research. 2018;61:863–905. doi:10.1613/jair.1.11192.

26. CodeBlue. Selangkah Covid-19 app wins Singapore AI Award for Health Tech; 2022. Available from: https://codeblue.galencentre.org/2022/04/29/selangkah-covid-19-app-wins-singapore-ai-award-for-health-tech/.

27. admin selangkah. Tutorial: Covid assessment centre (CAC) & home quarantine registration; 2021. Available from: https://selangkah.my/?p=11448.

28. Zhang H, Wu Y, He Y, Liu X, Liu M, Tang Y, et al. Age-Related Risk Factors and Complications of Patients With COVID-19: A Population-Based Retrospective Study. Frontiers in medicine. 2021;8.

29. Feature selection with the Boruta algorithm;. Available from: https://search.r-project.org/CRAN/refmans/Boruta/html/Boruta.html.

30. Bergstra J, Yamins D, Cox D. Making a science of model search: Hyperparameter optimization in hundreds of dimensions for vision architectures. In: International conference on machine learning. PMLR; 2013. p. 115–123.

31. CodeBlue. Over 2 million covid-19 care packages given to B40 families; 2022. Available from: https://codeblue.galencentre.org/2022/03/22/over-2-million-covid-19-care-packages-given-to-b40-families/.

